# Effects of universal masking on Massachusetts healthcare workers’ COVID-19 incidence

**DOI:** 10.1101/2020.08.09.20171173

**Authors:** Fan-Yun Lan, Costas A Christophi, Jane Buley, Eirini Iliaki, Lou Ann Bruno-Murtha, Assaad J. Sayah, Stefanos N. Kales

## Abstract

**Background:** Healthcare workers (HCWs) and other essential workers are at risk for occupational infection during the COVID-19 pandemic. Several infection control strategies have been implemented. Particularly, evidence shows that universal masking can mitigate COVID-19 infection, though existing research is limited by secular trend bias.

**Aims:** To investigate the effect of hospital universal masking on COVID-19 incidence among HCWs compared to the general community population.

**Methods:** We compared the 7-day averaged incidence rates between a Massachusetts (USA) healthcare system and Massachusetts residents statewide. The study period was from March 17 (the date of first incident case in the healthcare system) to May 6 (the date Massachusetts implemented public masking). The healthcare system implemented universal masking on March 26, we allotted a 5-day lag for effect onset, and peak COVID-19 incidence in Massachusetts was April 20. Thus, we categorized March 17-31 as the pre-intervention phase, April 1-20 the intervention phase, and April 21-May 6 the post-intervention phase. Temporal incidence trends (i.e. 7-day average slopes) were compared using standardized coefficients from linear regression models.

**Results:** The standardized coefficients were similar between the healthcare system and the state in both the pre- and post-intervention phases. During the intervention phase, the healthcare system’s epidemic slope became negative (standardized β: -0.68, 95% CI: –1.06 - -0.31), while Massachusetts’ slope remained positive (standardized β: 0.99, 95% CI: 0.94 – 1.05).

**Conclusions:** Universal masking at the healthcare system was associated with flattening the COVID-19 curve among HCWs, while the infection rate continued to rise in the surrounding community.

## Introduction

COVID-19 is an occupational risk for all essential workers, including healthcare workers (HCWs) who are essential to maintain health and hospital systems [1]. Evidence has shown HCWs are at risk for infection due to direct exposure to co-workers, patients and contaminated environments [2]. Accumulating research suggests that optimal infection control strategies, including the use of personal protective equipment, can minimize the risk of SARS-CoV-2 transmission to HCWs [3, 4]. Among now established hospital mitigation measures, universal masking is thought to be one of the most effective means to protect HCWs. However, existing evidence of universal masking efficacy is largely based on self-comparison results (i.e. comparing pre- and post-masking phases) [5] instead of comparing HCWs with a reference population, and thus cannot be free from secular trend bias. Therefore, we conducted this study to investigate the effect of universal masking policy in a Massachusetts (USA) healthcare system, using the surrounding statewide population as a comparison group.

## Methods

This time-series study examined the daily changes in COVID-19 incidence among 1) the Massachusetts statewide population and 2) the HCWs of a Massachusetts community healthcare system, which has been testing symptomatic employees since the initial outbreak. The statewide data were derived from Massachusetts Department of Public Health [6]. The HCW cohort of the healthcare system and related ethical statement are described in our previous paper [7]. We calculated 7-day moving averaged COVID-19 incidence to account for daily fluctuation. The date of an incident case was defined as the date when a HCW called the occupational health “hotline” for triage, and its’ reported date for the state.

We defined the study period to be from March 17, 2020, when the first case of the healthcare system was identified, to May 6, 2020. The study period was divided into three phases of interest: pre-intervention phase (March 17-31), intervention phase (April 1-20), and post-intervention phase (April 21-May 6). The phases’ date cut-offs were determined as follows. First, the healthcare system implemented universal masking on March 26 and we allowed five more days for the policy to take effect. The policy included securing N95s for all direct care staff managing confirmed or suspect COVID-19 patients, and providing procedure masks to all other staff. Second, April 20 is the day when Massachusetts reached the peak of COVID-19 incidence (Figure). Finally, Massachusetts implemented a statewide masking policy on May 6.

We built linear regression models to investigate the temporal trends for each cohort’s 7-day averaged incidence within each of the three phases. Standardized beta coefficients were calculated and presented to account for the difference in scale between the healthcare system and the state. We further obtained the standard errors and 95% confidence intervals (95% CIs) of the standardized coefficients. The analyses were performed using R software (version 3.6.3) and SAS software (version 9.4, SAS Institute).

## Results

During the study period, incident cases in the healthcare system and the state were 142 and 75,493, respectively. In the pre-intervention phase, both the healthcare system and the state had strong increasing trends in the 7-day averaged COVID-19 incidence (Figure and Table) with overlapping slopes (0.96 (0.80– 1.13) and 0.99 (0.92– 1.07), respectively). While the temporal trend among Massachusetts residents kept increasing with a similar slope in the second phase (the intervention phase) (0.99 (0.94– 1.05)), that of the healthcare system decreased (−0.68 (−1.06– −0.31). In the post-intervention phase, following the epidemic peak, incidence among both populations showed overlapping negative slopes (−0.90 (−1.19– −0.60) and −0.99 (−1.07– −0.92)) (Figure and Table).

## Discussion

Our present study provides additional, independent evidence of the protective effect of universal masking in healthcare settings. The healthcare system’s epidemic curve was flattened, and in fact, demonstrated a decreasing daily incidence trend after implementing the masking policy, while the statewide infection rate continued to increase during the same time period. The findings are in agreement with a recently published study using HCW self-comparison [5]. In that study, the authors found a significant decrement in the SARS-CoV-2 positivity rate after the implementation of universal masking. We further confirm this effect by including a comparison group, eliminating potential secular trend bias that limits before- and after-time-series studies.

Evidence has shown that the use of personal protective equipment (PPE) can prevent HCWs from being infected [8]. Two epidemiological studies conducted in Wuhan, China investigated 278 and 420 frontline HCWs with optimal PPE protection from different healthcare systems, respectively, and reported none infected [3, 9]. On the other hand, in the one study, 10 out of 213 HCWs without appropriate masking protection sustained nosocomial infection [9]. Furthermore, a Singaporean research team examined the surfaces of used PPE from HCWs who took care of a symptomatic COVID-19 patient and failed to detect SARS-CoV-2 (the virus causing COVID-19) from the collected samples [10]. In accordance with the existing evidence, our results further support masking’s protective effects from the perspective of hospital infection prevention.

The current study has several strengths such as the use of a comparable reference group, a validated outcome measurement, and a distinct intervention being implemented. Nonetheless, there are also some limitations. First, the current study is limited by small sample size of HCWs, the short intervention period and individual HCW compliance with the masking policy was not measured. Larger-scale studies are warranted. Second, the five-day interval assumed for the policy to take effect was somewhat arbitrary, but reasonable since most people develop symptoms within five days of infection. Finally, there could be unmeasured confounding, such as improved practices regarding social distancing, hand hygiene, and isolation of COVID-19 patients. However, our results show significant differences between the estimates, with a drastic change from positive to negative slope shortly after the masking policy’s implementation, which are unlikely to be entirely biased.

In conclusion, our results suggest a significant mitigation effect of universal masking on COVID-19 incidence when implemented in the healthcare setting, which may be applicable to other essential workers and indoor businesses.

### Key points

- **What is already known about this subject**
  - Healthcare workers (HCWs) are occupationally exposed to SARS-CoV-2.
  - Protecting HCWs and other essential workers is important to maintain healthcare and other essential businesses.
  - Optimal personal protective equipment (PPE) use can minimize workers’ risk of being affected by COVID-19.
- **What this study adds**
  - Additional evidence that universal masking is associated with decreased incident COVID-19 infections among HCWs.
  - Universal masking in the healthcare setting protects HCWs.
- **What impact this may have on practice or policy**
  - Timely universal masking policy should be implemented and maintained in healthcare settings during the COVID-19 pandemic.
  - Our study indirectly supports universal masking by other essential workers and indoor businesses.

## Data Availability

De-identified data are available upon request.

https://www.mass.gov/info-details/covid-19-response-reporting

---

**Figure.**
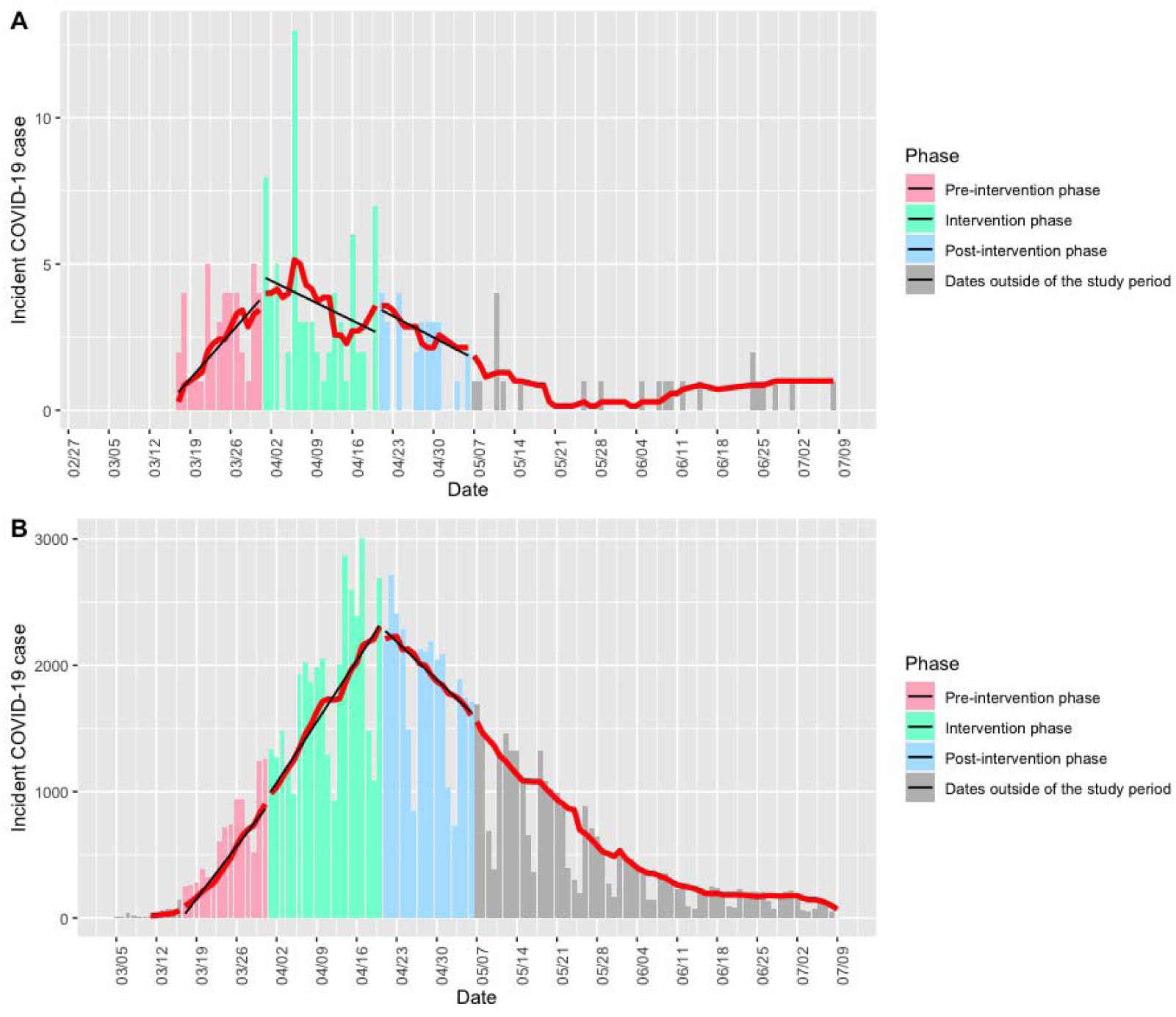
Temporal distribution of COVID-19 incidence among the (A) healthcare system employees and (B) Massachusetts residents statewide. Bars indicate the absolute number of daily cases. The red line denotes the 7-day average of new cases. The black lines show linear regression slopes in each specific phase. The phases were categorized based on the implementation date of universal masking policy by the healthcare system (pre-intervention phase, March 17-31; intervention phase, April 1-20; post-intervention phase, April 21-May 6).

**Table.** The crude and standardized beta coefficients of 7-day averaged COVID-19 incident cases regressed on days during the three study phases.

|  | Pre-intervention phase<br>(March 17-31) | Intervention phase<br>(April 1-20) | Post-intervention phase<br>(April 21-May 6) |
| --- | --- | --- | --- |
| The healthcare system |  |  |  |
| Crude | 0.26 (0.19– 0.26) | -0.10 (-0.15– -0.04) | -0.10 (-0.14– -0.07) |
| Standardized | 0.96 (0.80– 1.13) | -0.68 (-1.06– -0.31) | -0.90 (-1.19– -0.60) |
| Massachusetts statewide |  |  |  |
| Crude | 59.25 (54.73– 63.77) | 69.14 (65.31– 72.97) | -42.90 (-46.05– -39.76) |
| Standardized | 0.99 (0.92– 1.07) | 0.99 (0.94– 1.05) | -0.99 (-1.07– -0.92) |

Beta coefficient (95% CI) derived from linear regression models in each phase.

